# Response to antibiotic treatment of bacterial vaginosis predicts the effectiveness of LACTIN-V (*Lactobacillus crispatus* CTV-05) in the prevention of recurrent disease

**DOI:** 10.1101/2023.10.19.23297283

**Authors:** Anke Hemmerling, Michael R. Wierzbicki, Eric Armstrong, Craig R. Cohen

## Abstract

**Objectives:** Live biotherapeutic products (LBPs) containing vaginal *Lactobacillus crispatus* are promising adjuvant treatments to prevent recurrent bacterial vaginosis (BV) but may depend on the success of initial antibiotic treatment.

**Methods:** A *post hoc* analysis of data collected during the phase 2b LACTIN-V randomized control trial (*L. crispatus* CTV-05) explored the impact of clinical BV cure defined as Amsel criteria 0 of 3 (excluding pH, per 2019 FDA guidance) two days after completion of treatment with vaginal metronidazole gel on the effectiveness of an 11-week LACTIN-V dosing regimen to prevent BV recurrence by 12 and 24 weeks.

**Results:** At enrollment, 88% of participants had achieved clinical BV cure. The effect of LACTIN-V on BV recurrence compared to placebo differed by initial clinical BV cure status (p=0.02 by 12 weeks, and p=0.08 by 24 weeks). The LACTIN-V to placebo risk ratio of BV recurrence by 12 weeks was 0.56 (CI: 0.35, 0.77) among participants with initial clinical BV cure, and 1.34 (CI: 0.47, 2.23) among participants without clinical BV cure. Among women receiving LACTIN-V, those who had achieved clinical BV cure at enrollment reached higher levels of detectable *L. crispatus* CTV-05 compared to women failing to achieve clinical BV cure.

**Conclusion:** LACTIN-V appears to only decrease BV recurrence in women with clinical cure of BV following antibiotic treatment. Future trials of LBPs should consider limiting enrollment to these women.

## INTRODUCTION

Bacterial vaginosis (BV) is a genital tract infection in women characterized by a highly diverse vaginal microbiome and few lactobacilli. BV is associated with an elevated risk of adverse reproductive health outcomes such as preterm birth and the acquisition of sexually transmitted infections (STIs) including HIV. For decades, treatment has been limited to antibiotics, but BV recurrence is common.^1,2^ Clinical research has explored the use of live biotherapeutic products (LBPs) after antibiotics to optimize the vaginal microbiome by replenishing naturally occurring *Lactobacillus* strains.^3^ In the phase 2b trial of LACTIN-V (*Lactobacillus crispatus* CTV-05), we identified a significant reduction in BV recurrence by 12 weeks (one week after LACTIN-V or placebo) (RR 0.66; CI 0.44 to 0.87) and 24 weeks (three months after LACTIN-V or placebo) (RR 0.73; 95% CI 0.54 to 0.92) compared to placebo.

By 12 weeks, 30% of participants receiving LACTIN-V and 45% receiving placebo had experienced BV recurrence. Among women receiving LACTIN-V, 79% successfully colonized with *L. crispatus* CTV-05 at week 12, and those colonized had a median concentration of 1.7×10^6^ CFU/ml.^4^

We wanted to assess if the higher-than-expected BV recurrence in the LACTIN-V group could be associated with a lack of clinical BV cure following initial antibiotic treatment. Thus, we conducted this *post hoc* analysis to determine whether the effect of LACTIN-V on 1) the risk of BV recurrence and on 2) vaginal colonization of *L. crispatus* CTV-05 at 12- and 24-weeks differed by clinical BV cure status at enrollment.

## MATERIALS AND METHODS

In the phase 2b study^4^, 228 women aged 18-45 years diagnosed with BV were treated with 5 days of vaginally administered metronidazole 0.75%. Women with at least three Amsel criteria on wet mount and a Nugent score of 4-10 on gram stain at screening returned for enrollment within 48 hours after completing metronidazole. Women were randomized 2:1 to receive LACTIN-V or placebo over 11 weeks, followed by a 13-week post-dosing phase. Detection and quantification of *L. crispatus* (all strains, and the CTV-05 strain specifically) was measured by quantitative polymerase-chain-reaction (PCR) assays.

For this *post hoc* analysis, we followed the current 2019 Food & Drug Agency (FDA) guidance^5^ and performed a clinical test of cure (TOC) for BV at enrollment, seven days after initiating the five-day course of antibiotics. We used the following definitions for TOC status at enrollment, and for recurrent BV at follow-up visits:

1. Clinical TOC status at Enrollment: Based on wet mount. Clinical BV Cure determined as Amsel criteria 0 out of 3 (excluding pH), per 2019 FDA guidance.^5^
2. BV diagnosis at screening and recurrent BV after enrollment was the combination measure of Amsel criteria ≥ 3 of 4 (including pH) <u>and</u> a Nugent score of 4-10. This was based on the FDA Draft Guidance on Metronidazole from April 2013^6^, and a review of conducted BV studies while planning the phase 2b study.

To explore whether the effect of LACTIN-V differed by clinical TOC status after antibiotics at enrollment, the relative risk of BV recurrence was estimated via a log-binomial model with covariates for treatment group and baseline clinical TOC status along with an interaction term. Like the phase 2b analysis^4^, the models were performed in the ITT population and imputations of missing/unknown BV recurrence statuses were performed using logistic-regression multiple imputation under the assumption of a monotone missing data pattern. The relative risk estimates and their confidence intervals (CIs) at both levels of clinical TOC status (cure/failure) were obtained from the interaction model.

For both the log-binomial and proportional hazards models, separate models were run for the 12-week and 24-week time points. Analyses excluded the single participant in the LACTIN-V arm that did not receive study product and did not have wet mount results at enrollment available. All other randomized participants had determinable TOC status at enrollment and were included in recurrence analyses. For women receiving LACTIN-V, the colonization risk ratio comparing clinical BV cure to clinical failure to achieve BV cure at enrollment among LACTIN-V recipients was estimated using a log-binomial model with clinical status as a covariate. Missing values for *L. crispatus* CTV-05 concentration data were not imputed.

## RESULTS

### Test of BV cure after antibiotic treatment

Of the women receiving study product, a similar proportion of participants achieved clinical BV cure at enrollment among those randomized to LACTIN-V (134 of 151 [88.7%]) and placebo (67 of 76 [88.2%]). Sociodemographic data were similar between the ITT population (n=228) and those in the clinical BV cure subset (n=201) (data not shown).

### Recurrence of BV

By 12 weeks in the clinical cure cohort, 34 of 134 (25.4%) receiving LACTIN-V and 29 of 67 (43.3%) receiving placebo experienced recurrent BV (relative risk (RR) = 0.56; 95% CI: 0.35 to 0.77). In the clinical failure cohort, 12 of 17 (70.6%) and 5 of 9 (55.6%) had recurrent BV diagnosed by 12 weeks in the LACTIN-V and placebo groups, respectively (RR = 1.34; 95% CI: 0.47 to 2.23) (Figure 1). By 12 weeks, the difference in treatment effect between the clinical cure and failure cohorts was statistically significant (p = 0.02). Similarly, BV recurrence by 24 weeks in the clinical cure cohort was observed among 46 of 134 (34.3%) in the LACTIN-V group and 35 of 67 (52.2%) in the placebo group (relative risk (RR) = 0.67; 95% CI: 0.48 to 0.87). In the clinical failure cohort, 13 of 17 (76.5%) in the LACTIN-V group and 6 of 9 (66.7%) in the placebo group experienced recurrent BV by the 24-week visit (RR = 1.12; 95% CI: 0.57 to 1.68), with an interaction p-value of 0.08 (Figure 1).

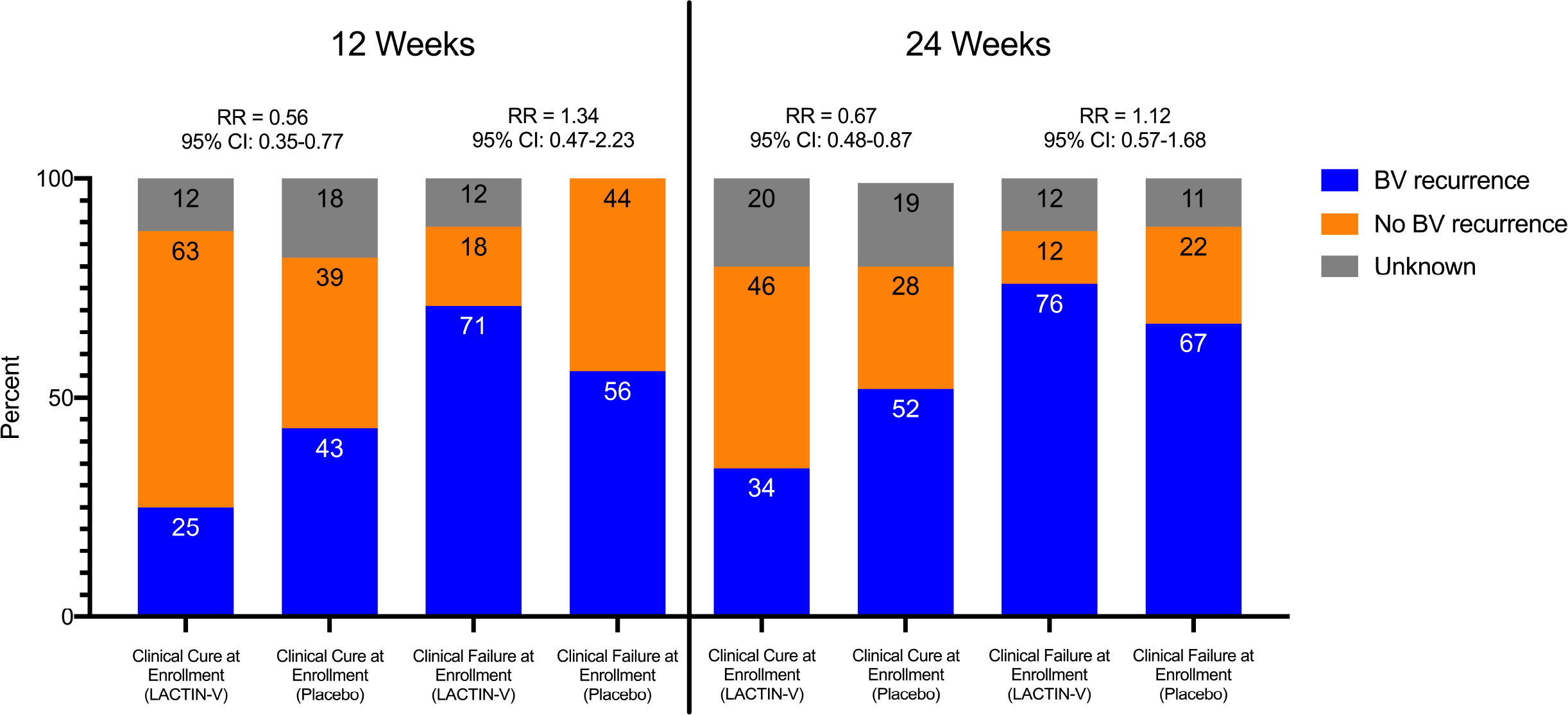

### Colonization with *L. crispatus* CTV-05

Among women receiving LACTIN-V with clinical BV cure and failure at enrollment, 83.6% and 40.0% respectively, colonized with *L. crispatus* CTV-05 at the 12-week visit (RR = 2.09; 95% CI: 1.12 to 3.91). The median concentrations among those successfully colonized at week 12 within the BV cure and failure cohorts were 2.0 ×10^6^ and 9.7×10^4^ CFU/ml, respectively. Similarly at the 24-week timepoint, 51.9% of the clinical BV cure subset and 20.0% of the clinical BV failure subset were colonized with *L. crispatus* CTV-05 (RR = 2.59; 95% CI: 0.93 to 7.26).

## DISCUSSION

### What this study adds

Most women achieved clinical BV cure within 7 days of initiating metronidazole and a sub-analysis confirmed that metronidazole quickly depleted BV-associated bacteria^7^. However, over half of the women randomized to placebo experienced BV recurrence by 24 weeks, confirming the limited sustained effect of antibiotic regimens alone.^1, 2^

While the safety and effectiveness of LACTIN-V to prevent recurrent BV was demonstrated previously^4^, this *post hoc* analysis shows that clinical cure of BV following antibiotic treatment is crucial to laying the groundwork for colonization with *L. crispatus* CTV-05, sustaining low rates of BV recurrence, and maximizing the clinical benefits of LACTIN-V.

### Implications for practice and research recommendations

Based on our findings, future studies that evaluate LBP-based treatment for BV should consider excluding women who fail to achieve clinical BV cure after initial antibiotic treatment. Our results also support the use of wet mount to evaluate the presence of clue cells and confirm clinical cure of BV soon after antibiotic treatment. Previous work has shown that there is a minimal expansion of lactobacilli shortly after antibiotic treatment for BV^7^, so it follows that testing for clinical cure should assess the absence of BV-associated bacteria, rather than the presence of lactobacilli. The Gram stain, which is central to diagnosing BV by Nugent score and evaluates lactobacilli and BV-associated bacteria^8^, may be more useful when determining BV recurrence at a later point after antibiotics when recovery of lactobacilli is expected.

### Limitations

The LACTIN-V phase 2b trial enrolled an ethnically diverse cohort from the several US-based sites, but the applicability of our findings to other populations may be limited. For example, the potential of *Lactobacillus*-based LBPs to reduce risk of HIV acquisition is particularly relevant for women in eastern and southern Africa, where HIV prevalence is high^9^. The composition of the vaginal microbiome tends to differ among women in this region which may have implications for optimizing *Lactobacillus*-based LBPs^9^. We were also limited in our ability to assess the relationship between BV cure and LACTIN-V efficacy from a multi-omic perspective. Future studies should explore potential mechanisms underpinning this relationship with specific focus on vaginal microbiome dynamics following antibiotics, metabolic determinants of *Lactobacillus* colonization, and transcriptional activity of *Lactobacillus* required for longer-term colonization.

## CONCLUSIONS

Our results indicate that clearing BV-associated microbes prior to *Lactobacillus*-based LBP administration may be crucial to maximizing their efficacy. A better understanding of the vaginal microbiome, metabolome, and metatranscriptome will allow a focused development of new tools to prevent BV, and ultimately help to prevent adverse reproductive health outcomes among women.

## KEY MESSAGES

- Topical metronidazole resolves most clinical BV in the near-term, though rapid recurrence is common.
- The effectiveness of LACTIN-V to prevent recurrent BV is higher in persons achieving clinical BV cure after initial antibiotic treatment.
- Colonization with *L. crispatus* CTV-05 is strongly correlated with clinical BV cure after initial antibiotic treatment.

## Data Availability

Data is not available due to the contractual nature of the funding from NIH.

